# Safety, tolerability, and immunogenicity of a SARS-CoV-2 recombinant spike protein vaccine: a randomised, double-blind, placebo-controlled, phase 1-2 clinical trial (ABDALA Study)

**DOI:** 10.1101/2021.11.30.21267047

**Authors:** Francisco Hernández-Bernal, Maria C. Ricardo-Cobas, Yenima Martín-Bauta, Zadis Navarro-Rodríguez, Marjoris Piñera-Martínez, Joel Quintana-Guerra, Karen Urrutia-Pérez, Klaudia Urrutia-Pérez, Cristina O. Chávez-Chong, Jorge L. Azor-Hernández, José L. Rodríguez-Reinoso, Leonardo Lobaina-Lambert, Elizabeth Colina-Ávila, Jacqueline Bizet-Almeida, Jeniffer Rodríguez-Nuviola, Sergio del Valle-Piñera, Mayara Ramírez-Domínguez, Elisangela Tablada-Ferreiro, Marel Alonso-Valdés, Gilda Lemos-Pérez, Gerardo E. Guillén-Nieto, Ariel Palenzuela-Díaz, Enrique Noa-Romero, Miladys Limonta-Fernández, Juan M. Fernández-Ávila, Nabil A. Ali-Mros, Lianne del Toro-Lahera, Rossana Remedios-Reyes, Marta Ayala-Ávila, Verena L. Muzio-González, for the ABDALA Group of Investigators

## Abstract

**Aim:** To evaluate the safety and immunogenicity of a SARS-CoV-2 recombinant spike protein vaccine (Abdala), administered intramuscularly in different strengths and vaccination schedules.

**Method:** A phase 1-2, randomized, double-blind, placebo-controlled trial was done. Subjects were randomly distributed in 3 groups: placebo, 25 and 50µg RBD. The product was applied intramuscularly, 0.5 mL in the deltoid region. During the first phase, two immunization schedules were studied: short (0-14-28 days) and long (0-28-56 days). In phase 2, only the short scheme was evaluated. The main endpoints were: safety and proportion of subjects with seroconversion of anti-RBD IgG antibodies to SARS-CoV-2. Blood samples were collected in several points according to the corresponding vaccination schedule to determine the level of RBD-specific IgG antibodies (seroconversion rates and geometric mean of the titers), the percentage of inhibition of RBD-ACE-2 binding and levels of neutralizing antibodies.

**Results:** The product was well tolerated. Severe adverse events were not reported. Adverse reactions were minimal, mostly mild and local (from the injection site), resolved in the first 24-48 hours without medication. In phase 1, at day 56 (28 days after the third dose of the short vaccination schedule, 0-14-28 days) seroconversion of anti-RBD IgG was seen in 95.2 % of the participants (20/21) for the 50μg group and 81 % of the participants (17/21) for the 25μg group, and none in the placebo group (0/22); whereas neutralizing antibodies to SARS-CoV-2 were seen in 80 % of the participants (8/10) for the 50μg group and 94.7% of the participants (18/19) for the 25μg group. For the long schedule, at day 70 (14 days after the third dose) seroconversion of anti-RBD IgG was seen in 100% of the participants (21/21) for the 50μg group and 94.7% of the participants (18/19) for the 25μg group, and none in the placebo group (0/22); whereas neutralizing antibodies to SARS-CoV-2 were seen in 95 % of the participants (19/20) for the 50μg group and 93.8% of the participants (15/16) for the 25μg group In phase 2, at day 56 seroconversion of anti-RBD IgG was seen in 89.2% of the participants (214/240) for the 50μg group, 77.7% of the participants (185/238) for the 25μg group, and 4.6% in the placebo group (11/239); whereas neutralizing antibodies to SARS-CoV-2 were seen in 97.3% of the participants (146/150) for the 50μg group and 95.1% of the participants (58/61) for the 25μg group.

**Conclusion:** Abdala vaccine against SARS-CoV-2 was safe, well tolerated and induced humoral immune responses against SARS-CoV-2 among adults from 19 to 80 years of age.

**Trial registration / Review protocol:** RPCEC00000346. Cuban Public Clinical Trial Registry (WHO accepted Primary Registry).

Available from: https://rpcec.sld.cu/en/trials/RPCEC00000346-En

**Information about the ethical aspects and IRB approval:** The protocol was approved by the Ethic Committee of the participating hospital and by the Cuban Regulatory Authority (Center for State Control of Drugs, Medical Devices and Equipment).

**Summary box:** COVID-19 is a serious global health problem. Vaccines are urgently needed to protect humanity. Multiple vaccine candidates are currently being evaluated. The article shows promising safety and immunogenicity results for a vaccine candidate, based on the recombinant RBD subunit of the spike protein.

## Introduction

The global pandemic of the new 2019 coronavirus disease (COVID-19) caused by the severe acute respiratory syndrome coronavirus 2 (SARS-CoV-2) started in Wuhan, China in December 2019, and from then on has spread throughout the world. [1, 2]

The clinical spectrum of a SARS–CoV-2 infection ranges from the absence of symptoms (asymptomatic infection) or mild respiratory symptoms to severe acute respiratory illness and death. Initially, it manifests mainly as fever, but sometimes only chills and respiratory symptoms occur due to mild dry cough and gradual dyspnea, in addition to fatigue and even diarrhea. In severe cases, the disease can progress rapidly, causing acute respiratory distress syndrome (ARDS), pneumonia, septic shock, irreversible metabolic acidosis, multiple organ failure, and clotting disorders, among other complications. The prognosis varies from recovery in most cases to torpid evolution and death. [2, 3]

Vaccines are urgently needed to mitigate the consequences of this pandemic and protect humanity from future epidemics caused by this virus. In this sense, clinical trials with multiple vaccine candidates, with accelerated designs and overlapping of the traditional phases of clinical research, are currently carried out worldwide, without breaching Good Clinical Practices (GCP). [4] Obtaining safe and effective preventive vaccines, as well as implementing them with broad global coverage, would be the fastest and safest strategy to manage this terrible pandemic. [5]

At the Centre for Genetic Engineering and Biotechnology in Havana, work has been made on several vaccine candidates using platforms already known to this institution and also considering the state-of-the-art of research around COVID-19, especially the immunological aspects necessary for the development of vaccines against this infection. One of these vaccine candidates was Abdala, based on the recombinant RBD subunit of the spike protein produced in *Pichia pastoris* yeast, [6] adjuvanted to alumina.

The aim of the present work was to evaluate the safety and immunogenicity of the Abdala vaccine, administered intramuscularly in different strengths and schedules, for specific active immunization against SARS-CoV-2 infection in adults between 19 and 80 years of age.

## Methods

A randomised, adaptive, double blind, placebo-controlled, phase 1-2 clinical trial was carried out in “Saturnino Lora” Hospital, Santiago de Cuba. Subjects aged between 19 and 54 years (for phase I) and between 19 and 80 years (for phase II), who gave their written, informed consent to participate, were eligible. Exclusion criteria were: virological diagnosis by RT-PCR of infection to SARS-CoV-2, contact or suspect of COVID-19, subjects at high risk of exposure to SARS-CoV-2 infection (health workers in 1st line of medical care), acute infection in the last 15 days, autoimmune or endocrine-metabolic diseases decompensated at the time of inclusion, subject treated in the last three months or with any medical condition that requires an immunomodulator, steroid (except topical or inhaled) or cytostatic during the study. Individuals with body mass index ≤18 or ≥35 Kg/m^2^, tattoos in both deltoid regions, administration of any research product in the last three months, allergy to thiomersal or any other component of the medicament, pregnancy or breastfeeding, and mental disorders, were also excluded.

The trial was conducted in medical wards and certified areas for the vaccination process. The participating researchers were specialists in internal medicine and intensive care, and the vaccination under study was administered by specialized nurses. The protocol followed the Declaration of Helsinki guidelines and was evaluated by the Ethics and Review Committee of the Provincial Hospital “Saturnino Lora” in Santiago de Cuba (clinical site participating in the trial), who granted ethical approval of the study. This Institutional Review Boards was made up of highly qualified medical specialists not linked to the study, as well as a member of the community. This committee followed up on the research ensuring the protection of the rights, safety and well-being of the subjects involved in the study. In addition, the Cuban Center for State Control of Drugs, Medical Devices and Equipment approved the start of the clinical trial after considering the scientific, methodological and ethical aspects.

Recombinant protein of the receptor-binding domain (RBD) of the SARS-CoV-2 virus (Abdala vaccine active ingredient) was produced in *Pichia pastoris* at the Center for Genetic Engineering and Biotechnology in Havana. It is a slightly opaque grayish-white suspension that separates, after a settling time, into two phases: one transparent liquid and the other in the form of a gel, which when shaken is easily resuspended and is essentially free of foreign particles. Each mL contains 50 or 100 µg RBD, 0.60 mg aluminum hydroxide, 0.56 mg disodium hydrogen phosphate, 0.62 mg sodium dihydrogen phosphate dihydrate, 8.5 mg sodium chloride and 0.05 mg thiomersal. Placebo vial had the same excipients, with the exception of the RBD additive. Their organoleptic characteristics and presentations were identical.

The subjects included were randomly distributed (1:1:1) to 3 groups: I) placebo; II) 25 µg RBD and III) 50 µg RBD. The product was applied intramuscularly, 0.5 mL in the deltoid region. During the first phase, two immunization schedules were studied: short (0-14-28 days) and long (0-28-56 days). In phase 2, only the short scheme (0-14-28 days), selected during the interim analysis, was evaluated. Concomitant treatment was not anticipated.

The main endpoints were: safety and proportion of subjects with seroconversion of anti-RBD IgG antibodies to SARS-CoV-2. The adverse events (type, duration, severity, outcome, and causality relationship) were carefully registered. The severity of the adverse events was classified in three levels: (a) mild, if no therapy was necessary; (b) moderate, if a specific treatment was needed, and (c) severe, when hospitalisation or its prolongation was required, the reaction was life-threatening or contributed to patient’s death. A qualitative assessment was used to classify the causal relationship as definite, probable, possible or doubtful. [7] Although it is a novel product, the RBD proteins obtained from different expression models have been evaluated in a large number of volunteers, with an adequate safety profile. Also, the adjuvant used has been widely used in humans. Adverse reactions associated with vaccination were especially sought (pain at the injection site, erythema, induration, headache, fever, among others). As part of the safety profile, hematological determinations (hemoglobin, hematocrit, hemogram with differential, platelet count) and blood chemistry (glycemia, cholesterol, creatinine, uric acid, transaminases) were performed at the beginning and 14 days after completing the scheme immunization (day 42 or 70, according to the short or long schedule, respectively).

In phase 1 trial, blood samples were collected in all groups at day 0, before the application of the first dose (baseline), and at several points according to the corresponding vaccination schedule (at days 42 and 56 for the schedule 0-14-28 days and at days 56 and 70 for the schedule 0-28-56 days) to determine the level of RBD-specific IgG antibodies, in terms of seroconversion rates and geometric mean of the titers (GMT), the percentage of inhibition of RBD-ACE-2 binding in terms of proportions and media (95 % CI) and levels of neutralizing antibodies to live SARS-CoV-2 in terms of proportions an GMT. In the phase 2 trial, blood samples were taken at time 0 and at days 42 and 56 (14 and 28 days after the third dose).

IgG antibodies were quantified by UMELISA SARS-CoV-2 anti-RBD (Immunoassay Centre, Havana). [8] Titers are given in arbitrary units per millilitre (AU/mL) with a cut-off value of 1.95. The percentage of inhibition to RBD-ACE-2 (angiotensin-converting enzyme 2) binding was determined using and in-house virus neutralization test (Center for Genetic Engineering and Biotechnology, Havana). Results are given in inhibition percentage. The assay threshold for positivity was 30%. The neutralizing antibodies titers were detected by a standard virus microneutralization assay using live SARS-CoV-2 (CUT2010-2025/Cuba/2020 strain) [9] carried out at Civilian Defense Scientific Research Center, Cuba. Viral neutralizing titers were calculated as the highest serum dilution with an optical density (OD) higher that the cut-off value (calculated as the average of the OD of the cell control wells divided by two).

Randomisation was carried out in the supply group of the Clinical Research Direction of the CIGB, in blocks of 12 or 6 individuals (phase 1 and 2, respectively), by means of a computerised random number generator. The site received the product in such blocks, in masked vials in order to prevent their identification, labelled with each subject’s number. Therefore, the decision to accept or reject a participant was made, and informed consent was obtained from the participant, in ignorance of the assignment in the sequence. All participants (investigators, subjects, and monitors) were kept blinded during all the study performance and data management. Statistical analyses were done without knowledge of the groups’ identity. This was known after the analyses were concluded.

In phase 1 of the trial, it was expected to identify the two best experimental groups which would continue to the second phase considering: a) the percentage of seroconversion of anti-RBD IgG antibodies to SARS-CoV-2; b) the non-occurrence of serious adverse events with a causal relationship attributable to the research product in no more than 5% of the subjects. During phase 2, a 50% superiority over the placebo group was expected in at least one of the selected experimental groups in terms of seroconverted subjects, without the occurrence of serious adverse events.

Assuming type I and II errors of 0.05 and 0.20, respectively, a sample size of 20 subjects was estimated using the PASS software (www.ncss.com). Considering 10% dropouts, the final sample size was 22 subjects per group (132 volunteers) for phase 1. Under the same assumptions, the final sample size during the second stage of the study was 242 individuals per group (726 subjects in total, including the 66 participants from phase I assigned to the short immunization schedule).

Statistical analyses were done with R version 3.6.2. The relationship between the qualitative response variables and the control variables was studied. To do this, contingency tables were constructed and the chi-square test was performed to analyse whether there were significant dependencies, and a correspondence analysis was carried out in the cases where this fact was verified. The immunogenicity analyses were done in the per-protocol population, refer to participants who fulfilled the inclusion and exclusion criteria, who completed the course of vaccination schedule and had valid immunogenicity results both before immunization and the indicated days after vaccination as defined in the study protocol. Individuals, who had anti-RBD IgG antibodies at the baseline time determination, before the first dose of the product was applied, were excluded from the immunogenicity analyses so that all study groups started from the same condition and the effect of the primary immunization schedule could be clearly assessed. We used the Pearson χ^2^ test for the analysis of categorical outcomes. We calculated 95% confidence intervals (CI) for all categorical outcomes using the Clopper-Pearson method. GMTs were calculated as the mean of the assay results after the logarithmic transformation was made; we then exponentiated the mean to express results on the original scale. Two-sided 95% CI were obtained by performing logarithmic transformations of titers, calculating the 95% confidence interval with reference to Student’s t-distribution, and then exponentiating the limits of the confidence intervals. We used the Kruskal-Wallis test as a non-parametric alternative for ANOVA to compare the log-transformed antibody titer. As a complement to this test, once the null hypothesis is rejected, multiple comparison tests with Bonferroni’s correction were performed to adjust by the number of comparisons. Hypothesis testing was two-sided and we considered p values of less than 0.05 to be significant. We used the Student’s t-test to compare the mean of two samples. Mann Whitney U test was used as a nonparametric alternative to Student’s t-test. Pearson’s linear correlations were also calculated to assess the association between responses on different assays by time of evaluation and study group. To assess safety, adverse events were tabulated and plotted by dose and study group.

An independent data monitoring committee consisted of one independent statistician, pathologist, clinician, epidemiologist and immunologist was established before commencement of the study. Safety data were assessed and reviewed by the committee to ensure the toxicity criteria of phase 1 were not met and allow the further proceeding of the clinical trial.

## Results

From December 2020 to April 2021 a total of 792 subjects were included out of 853 that were screened. Their disposition is shown in Figure 1. During phase 1, 132 subjects were included, randomly distributed into two vaccination schedules (0-14-28 and 0-28-56 days) and three study groups for each schedule (placebo and two RBD strengths: 25 and 50 μg). After an interim analysis of the results and given the complex epidemiological situation resulting from the COVID-19 pandemic, it was strategically decided to continue towards phase 2 of the trial with the three study groups of the short vaccination schedule (0-14-28 days). In this sense, phase 2 included 660 new individuals, to which were added the 66 subjects evaluated in a similar vaccination scheme during the first stage (726 subjects in total; 242 in each study group). All volunteers completed the vaccination schedule (three doses), except for five individuals assigned to the long schedule in phase 1 (2 in the placebo and RBD 25 g groups, and 1 in the 50 g group of the vaccine candidate) and four subjects during phase 2 (two in the placebo and lower strength vaccine groups).

**Figure 1.**
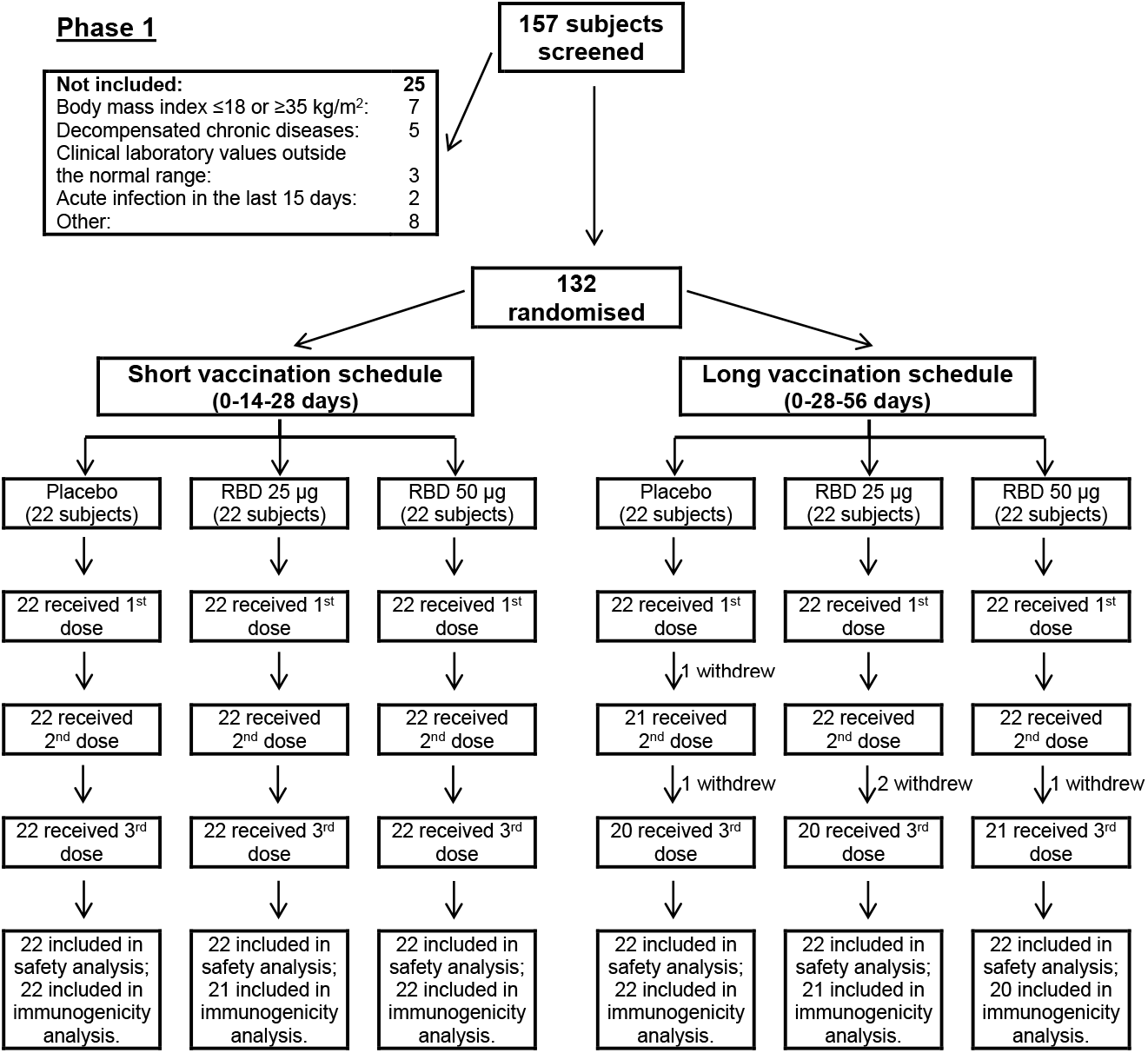
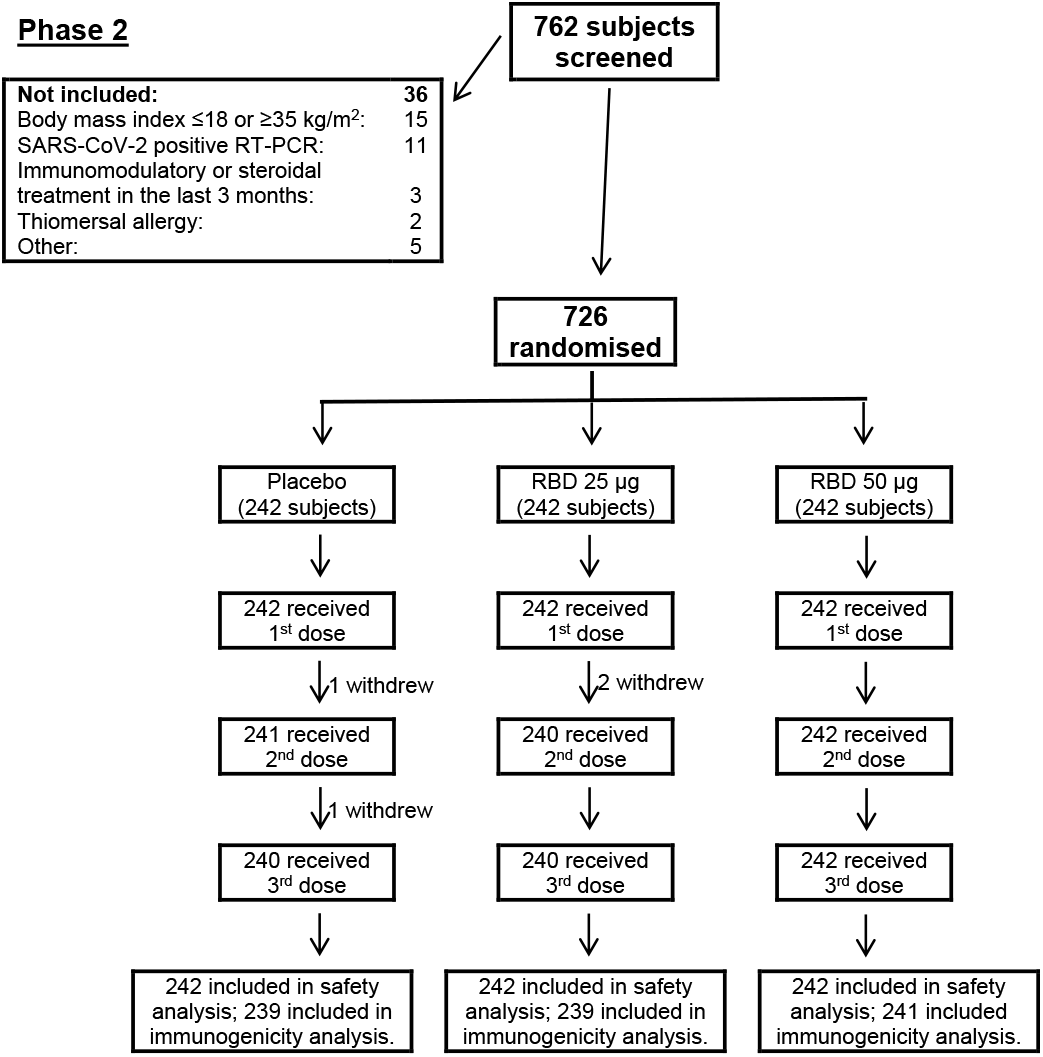
Disposition of 792 subjects from December 2020 to April 2021.

Table 1 shows the demographic and baseline characteristics of the subjects. Most of them were males, 19 to 80 years-old, with the ethnic distribution of the Cuban population in the south eastern region of the country. No relevant imbalances can be seen.

**Table 1.** Demographic characteristics of the participants in the Abdala trial at enrollment.

| Variable |  | Short schedule (0-14-28 days) |  |  |  | Long schedule (0-28-56 days) |  |  |  | Overall |
| --- | --- | --- | --- | --- | --- | --- | --- | --- | --- | --- |
|  |  | Placebo | RBD 25 µg | RBD 50 µg | Subtotal | Placebo | RBD 25 µg | RBD 50 µg | Subtotal |  |
| PHASE I (19-54 years) |  |  |  |  |  |  |  |  |  |  |
| N |  | 22 | 22 | 22 | 66 (50.0) | 22 | 22 | 22 | 66 (50.0) | 132 (100) |
| Sex – no. (%) | Female | 11 (50.0) | 7 (31.8) | 9 (40.9) | 27 (40.9) | 10 (45.5) | 12 (54.5) | 11 (50.0) | 33 (50.0) | 60 (45.5) |
|  | Male | 11 (50.0) | 15 (68.2) | 13 (59.1) | 39 (59.1) | 12 (54.5) | 10 (45.5) | 11 (50.0) | 33 (50.0) | 72 (55.5) |
| Age | years | 37.9 ± 10.0 | 40.7 ± 7.3 | 44.4 ± 9.2 | 41.0 ± 9.2 | 41.1 ± 9.4 | 39.9 ± 9.4 | 44.2 ± 8.3 | 41.7 ± 9.1 | 41.3 ± 9.1 |
| Ethnicity – no. (%) | White | 4 (18.2) | 4 (18.2) | 7 (31.8) | 15 (22.7) | 7 (31.8) | 3 (13.6) | 2 (9.1) | 12 (18.2) | 27 (20.5) |
|  | Black | 11 (50.0) | 2 (9.1) | 6 (27.3) | 19 (28.8) | 6 (27.3) | 4 (18.2) | 8 (36.4) | 18 (27.3) | 37 (28.0) |
|  | Mestizo | 7 (31.8) | 16 (72.7) | 9 (40.9) | 32 (48.5) | 9 (40.9) | 15 (68.2) | 12 (54.5) | 36 (54.5) | 68 (51.5) |
| BMI | Kg/m<sup>2</sup> | 25.2 ± 4.0 | 26.5 ± 4.7 | 25.5 ± 3.3 | 25.7 ± 4.0 | 24.4 ± 3.3 | 25.9 ± 3.3 | 26.3 ± 4.4 | 25.5 ± 3.7 | 25.6 ± 3.9 |
| PHASE II (19-80 years; short schedule: 0-14-28 days) |  |  |  |  |  |  |  |  |  |  |
| Age group: 19 - 54 years |  |  |  |  |  | Age group: 55 - 80 years |  |  |  |  |
| N |  | 153 | 149 | 151 | 453 (62.4) | 89 | 93 | 91 | 273 (37.6) | 726 (100) |
| Sex – no. (%) | Female | 76 (49.7) | 80 (53.7) | 76 (50.3) | 232 (51.2) | 28 (31.5) | 40 (43.0) | 37 (40.7) | 105 (38.5) | 337 (46.4) |
|  | Male | 77 (50.3) | 69 (46.3) | 75 (49.7) | 221 (48.8) | 61 (68.5) | 53 (57.0) | 54 (59.3) | 168 (61.5) | 389 (53.6) |
| Age | years | 35.7 ± 12.0 | 35.6 ± 11.4 | 30.1 ± 12.5 | 35.0 ± 12.0 | 65.7 ± 8.0 | 63.2 ± 7.5 | 64.0 ± 7.3 | 64.3 ± 7.2 | 46.5 ± 17.3 |
| Ethnicity – no. (%) | White | 53 (34.6) | 63 (42.3) | 51 (33.8) | 167 (36.9) | 28 (31.5) | 24 (25.8) | 35 (38.4) | 87 (31.9) | 254 (35.0) |
|  | Black | 34 (22.2) | 20 (13.4) | 17 (11.3) | 71 (15.7) | 25 (28.1) | 17 (18.3) | 16 (17.6) | 58 (21.2) | 129 (17.8) |
|  | Mestizo | 66 (43.1) | 66 (44.3) | 83 (55.0) | 215 (47.5) | 36 (40.4) | 52 (55.9) | 40 (44.0) | 128 (46.9) | 343 (47.2) |
| BMI | Kg/m<sup>2</sup> | 25.1 ± 4.2 | 25.2 ± 4.1 | 24.7 ± 4.1 | 25.0 ± 4.1 | 25.9 ± 4.0 | 27.2 ± 4.4 | 26.6 ± 4.4 | 26.6 ± 4.3 | 25.6 ± 4.3 |

Plus-minus values are means ± SD.
BMI: Body-mass index (is the weight in kilograms divided by the square of the height in meters. The calculation was based on the weight and height measured at the time of screening).

The product was well tolerated. Severe adverse events or adverse events of special interest were not reported and there were no withdrawals for this cause. In the phase 1 trial, the overall incidence of adverse reactions was 6/22 (27.3%) participants in the 25 and 50 μg groups, respectively, and 3/22 (13.6%) in the placebo group in the short vaccination schedule (0-14-28 days); and 8/22 (36.4%) in the 25 μg group, 9/22 (40.9%) in the 50 μg group, and 4/22 (18.2%) in the placebo group in the long vaccination schedule (0-28-56 days). During phase 2, adverse reactions were reported by 53/242 (21.9%) subjects in the 25 μg group, 75/242 (31.0%) in the 50 μg group, and 41/242 (16.9%) in the placebo group, all under the short vaccination schedule.

In the two phases of the clinical trial, overall reactogenicity was largely absent or mild in most reports, and the following doses were neither withheld nor delayed due to reactogenicity (Figure 2). After the first vaccination, local and systemic reactogenicity it was absent or decreased with the application of subsequent doses. Most of the adverse reactions resolved spontaneously in the first 24-48 hours without medication. There were no clinically relevant laboratory abnormalities associated with vaccination.

**Figure 2.**
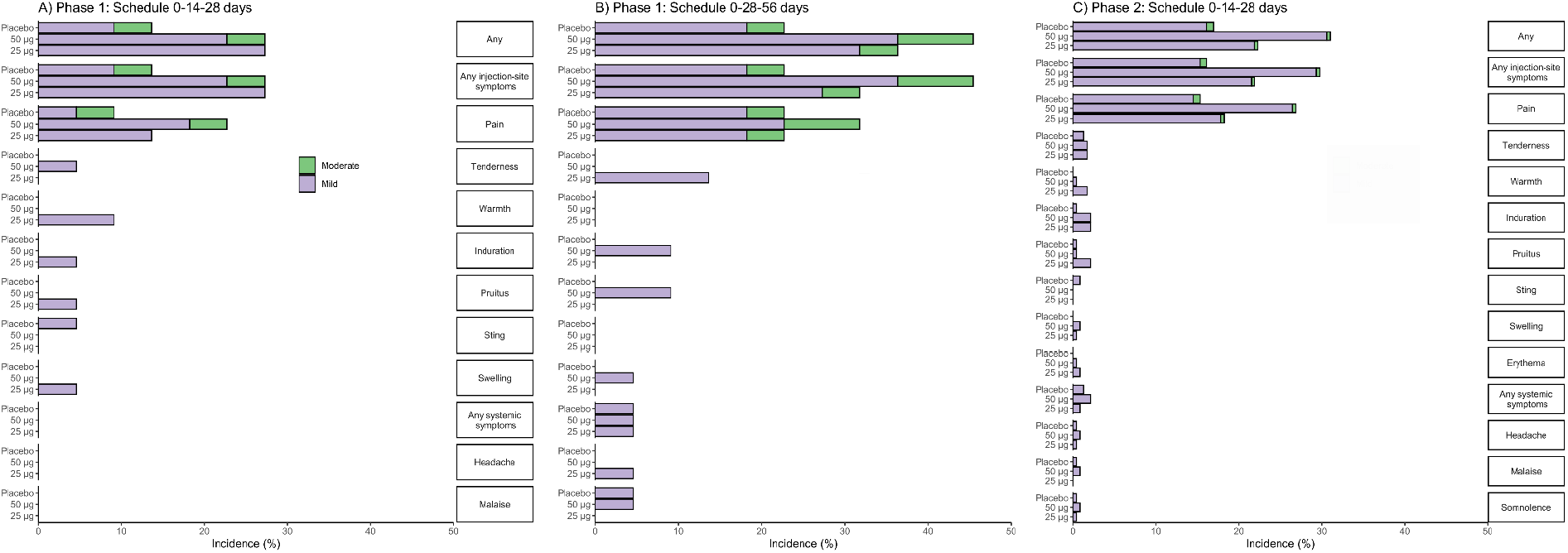
The percentage of participants in each study group (RBD 25 µg, RBD 50 µg, Placebo) with adverse reactions according to the maximum FDA toxicity grade (mild or moderate) from first dose up to 14 days after third dose is plotted by signs or symptoms. Participants who reported 0 events make up the remainder of the 100%.

The primary immunogenicity outcome was the seroconversion rate of the IgG RBD-binding antibodies. Seroconversion was defined as at least a four-fold increase of antibody titers over baseline. Secondary immunogenic endpoints were the magnitude of this response, in geometric (GMTs), percentage of inhibition to RBD-ACE-2 binding as well as the neutralizing antibodies to live SARS-CoV-2. In phase 2, antibody titers of the same groups from phase 1 trial were combined and analyzed together.

In phase 1, seroconversion rates of anti-RBD IgG for the short schedule (0-14-21 days), measured 14 days after the third dose (day 42), were 81% for the 25 μg group and 95.5% for the 50 μg group (p=0.19). At day 56 (28 days after the third dose) there were 81% and 95.2% for the 25 μg and 50 μg group respectively (p=0.21). The results for the long schedule (0-28-56 days) at day 56 (14 days after the application of the second dose) were 55 for the 25 μg group and 57.1% for the 50 μg group (p=1.0) that increased at day 70 (28 days after the third dose) to 94.7% and 100% for the 25 μg and 50 μg group (p=0.96) (Table 2).

**Table 2.**
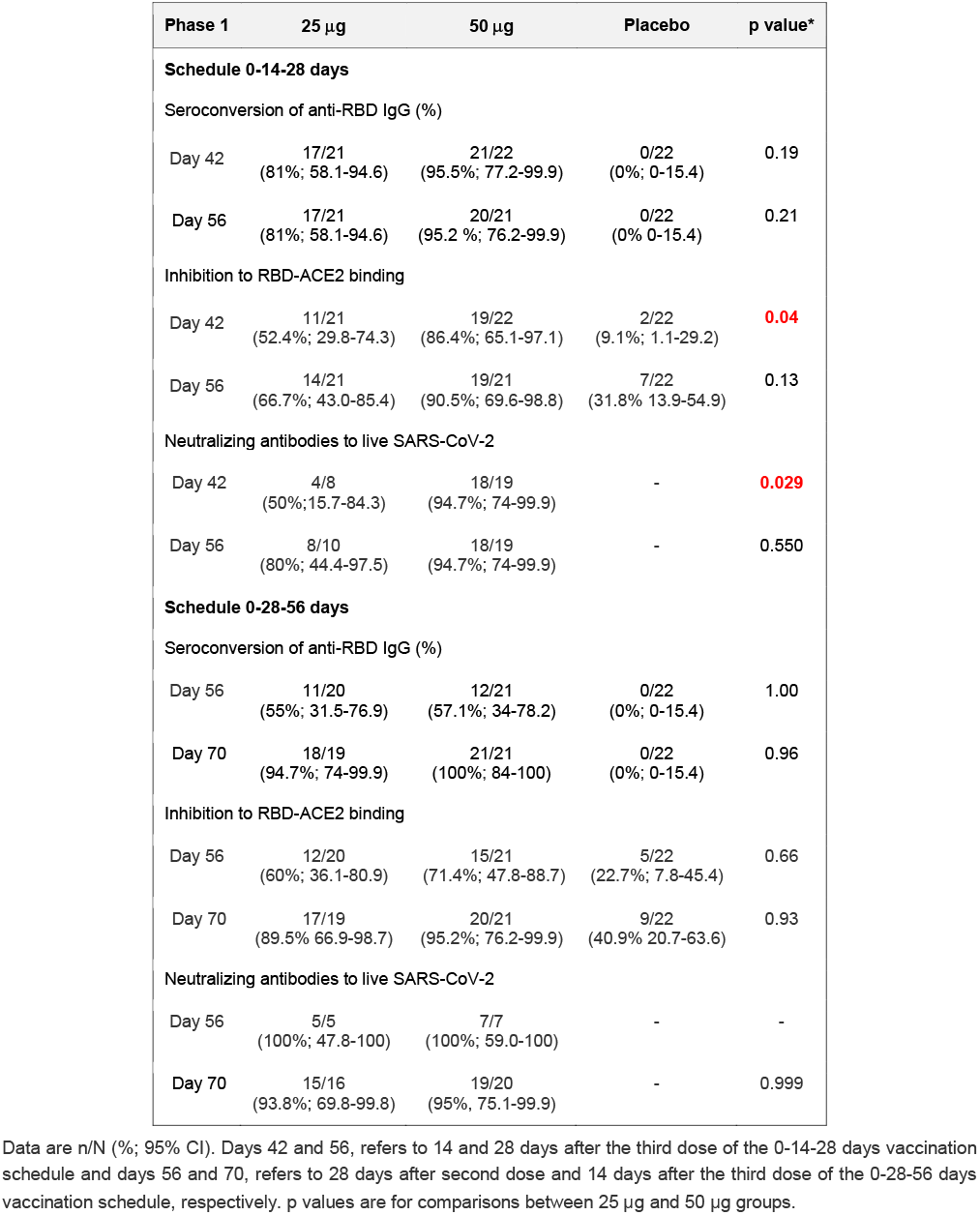
Seroconversion rates for anti-RBD IgG and proportion of individuals with inhibition to RBD-ACE2 binding and neutralizing antibodies to SARS-CoV-2.

| Phase 1 | 25 µg | 50 µg | Placebo | p value* |
| --- | --- | --- | --- | --- |
| **Schedule 0-14-28 days** |  |  |  |  |
| Seroconversion of anti-RBD IgG (%) |  |  |  |  |
| Day 42 | 17/21<br>(81%; 58.1-94.6) | 21/22<br>(95.5%; 77.2-99.9) | 0/22<br>(0%; 0-15.4) | 0.19 |
| Day 56 | 17/21<br>(81%; 58.1-94.6) | 20/21<br>(95.2 %; 76.2-99.9) | 0/22<br>(0% 0-15.4) | 0.21 |
| Inhibition to RBD-ACE2 binding |  |  |  |  |
| Day 42 | 11/21<br>(52.4%; 29.8-74.3) | 19/22<br>(86.4%; 65.1-97.1) | 2/22<br>(9.1%; 1.1-29.2) | **0.04** |
| Day 56 | 14/21<br>(66.7%; 43.0-85.4) | 19/21<br>(90.5%; 69.6-98.8) | 7/22<br>(31.8% 13.9-54.9) | 0.13 |
| Neutralizing antibodies to live SARS-CoV-2 |  |  |  |  |
| Day 42 | 4/8<br>(50%;15.7-84.3) | 18/19<br>(94.7%; 74-99.9) | - | **0.029** |
| Day 56 | 8/10<br>(80%; 44.4-97.5) | 18/19<br>(94.7%; 74-99.9) | - | 0.550 |
| **Schedule 0-28-56 days** |  |  |  |  |
| Seroconversion of anti-RBD IgG (%) |  |  |  |  |
| Day 56 | 11/20<br>(55%; 31.5-76.9) | 12/21<br>(57.1%; 34-78.2) | 0/22<br>(0%; 0-15.4) | 1.00 |
| Day 70 | 18/19<br>(94.7%; 74-99.9) | 21/21<br>(100%; 84-100) | 0/22<br>(0%; 0-15.4) | 0.96 |
| Inhibition to RBD-ACE2 binding |  |  |  |  |
| Day 56 | 12/20<br>(60%; 36.1-80.9) | 15/21<br>(71.4%; 47.8-88.7) | 5/22<br>(22.7%; 7.8-45.4) | 0.66 |
| Day 70 | 17/19<br>(89.5% 66.9-98.7) | 20/21<br>(95.2%; 76.2-99.9) | 9/22<br>(40.9% 20.7-63.6) | 0.93 |
| Neutralizing antibodies to live SARS-CoV-2 |  |  |  |  |
| Day 56 | 5/5<br>(100%; 47.8-100) | 7/7<br>(100%; 59.0-100) | - | - |
| Day 70 | 15/16<br>(93.8%; 69.8-99.8) | 19/20<br>(95%, 75.1-99.9) | - | 0.999 |

Data are n/N (%; 95% CI). Days 42 and 56, refers to 14 and 28 days after the third dose of the 0-14-28 days vaccination schedule and days 56 and 70, refers to 28 days after second dose and 14 days after the third dose of the 0-28-56 days vaccination schedule, respectively. p values are for comparisons between 25 µg and 50 µg groups.

None of the participants in the placebo groups seroconverted. For both strengths of RBD/vaccination schedules high seroconversion rates were obtained. At baseline, GMTs of anti-RBD IgG in all the participants were at the lower limit of quantitation (1.95). For the short schedule, by day 42, GMT had increased to 22.35 (95% CI 13.07-38.21) for the 25 μg group and to 131.20 (95% CI 70.65-243.61) for the 50 μg group. For the evaluation at day 56, GMT for the 25 μg group were 22.23 (95% CI 13.09-37.35) and GMT had further increased to 155.18 (95% CI 84.12-286.27) for the 50 μg group. Higher GMTs were found at 42 and 56 days for the 50 μg group, with significant differences between GMTs of 25 μg and 50 μg groups (p<0.0001) (Figure 3A). In the placebo groups GMTs remained as in the baseline evaluation and significant differences with both 25 μg and 50 μg groups were found (Figure 3A). For the long schedule, the results after only two administered doses (day 56) were 12.21 (95% CI 5.99-24.90) for the 25 μg group and 16.11 (95% CI 7.30-35.56) for the 50 μg group. Fourteen days after the full immunization schedule, GMT had further increased for both dose levels, 72.99 (95% CI 39.84-133.73) for the 25 μg group and 221.43 (95% CI 127.56-384.37) for the 50 μg group (p=0.02) (Figure 3B). Interestingly, at day 56 no statistical difference between 25 μg and 50 μg but this was found on day 70, after the full series of three doses. Once again, for both 25 μg and 50 μg groups, statistical differences with placebo were found at 56 and 70 days for the 50 μg group (Figure 3B).

**Figure 3.**
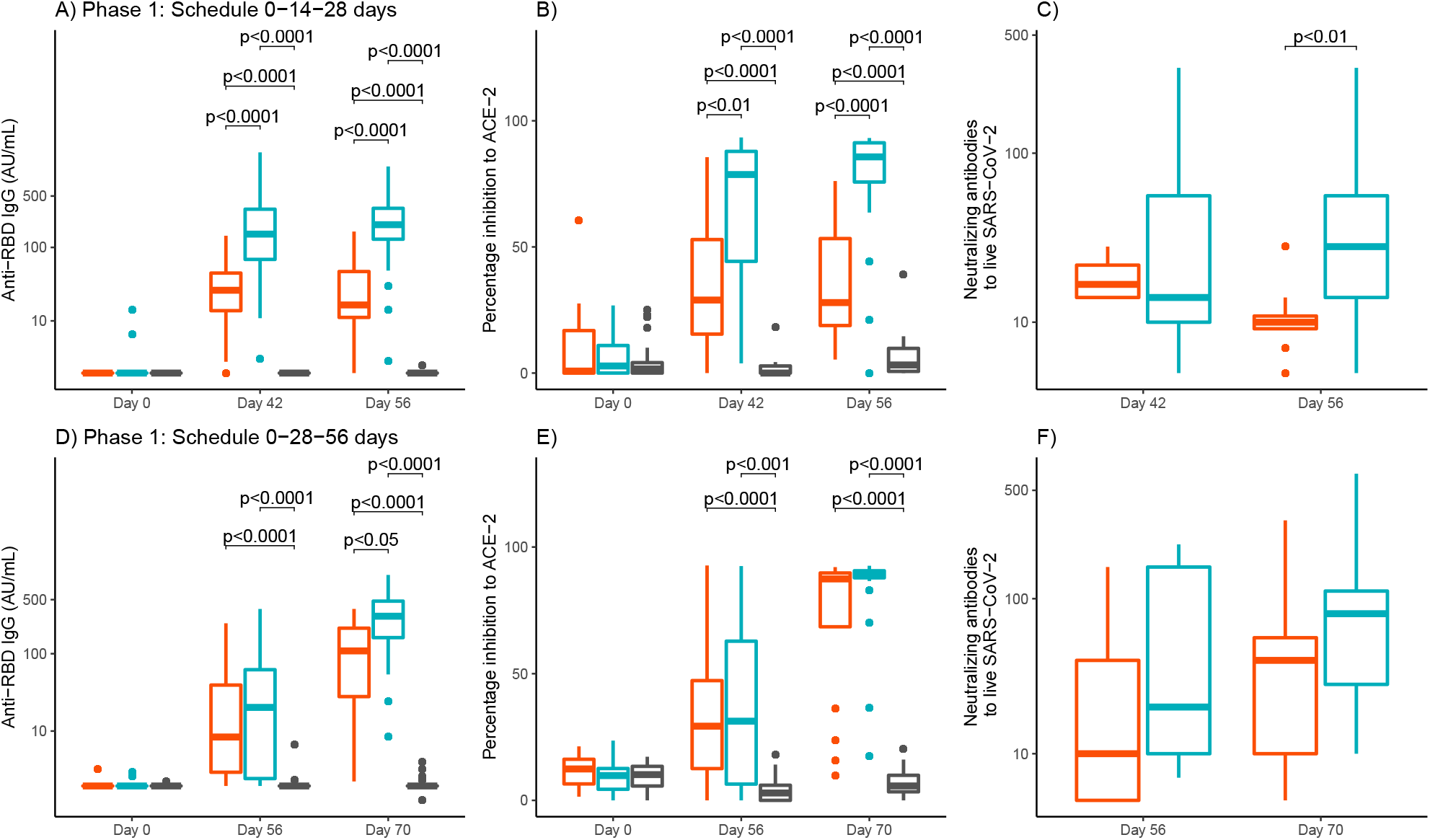
Quantitative variables of immunogenicity in the three study groups by schedule and days. From left to right are anti-RBD IgG antibody titers (graphs A and D), percentage of inhibition RDD-ACE2 binding (graphs B and E) and neutralizing antibody titers (graphs C and F). Study groups are represented by colors, red for RBD 25 µg, blue for RBD 50 µg and gray for placebo. The boxes and horizontal bars indicate interquartile range (IQR) and median, respectively. The whisker’s end points are the maximum and minimum values below or above the median ± 1.5 times the IQR. Points represent possible outliers. The braces contain the results of the Mann Whitney U multiple comparison tests with Bonferroni correction in cases where more than two groups appear (graphs A,B,D,E), and the Mann Whitney U tests in cases where only two groups appear (graphs C and F). Only p values for significant differences are shown.

The proportion of individuals with positive inhibition of RBD-ACE-2 binding for the short schedule at day 42 was 52.4% for the 25 μg group and 86.5% for the 50 μg group (p=0.04), increasing to 66.7% and 90.5% at day 56 for the corresponding (p=0.13) (Table 2). The media of the percentage of inhibition of RBD-ACE-2 binding for the short schedule at baseline were 8.83% (95% CI 1.92-15.73) for the 25 μg group and 5.73% (95% CI 2.39-9.07) for the 50 μg group, with no differences among the groups including placebo (Figure 3B). At day 42, the media had increased for both dose levels, for 25 μg group was 36.48% (95% CI 24.40-48.55) and 65.84% (95% CI 52.44-79.23) for the 50 μg group. By day 56, the results were 36.60% (95% CI 25.43-47.77) for the 25 μg group and a further increase was observed for the 50 μg group, 75.71% (95% CI 64.67-86.74). For both 42 and 56 days significant differences were obtained among the three groups of study, with highest media values for 50 μg group (Figure 3B). For the long schedule, the baseline results were 11.65% (95% CI 8.83-14.47) for the 25 μg group and 9.38% (95% CI 6.29-12.48) for the 50 μg group. At day 56, after only two applied doses, the percentage if inhibition had increased for both dose levels, 34.28% (95% CI 22.65-45.90) for the 25 μg group and 37.56% (95% CI 23.14-51.97) for the 50 μg group, with no significant differences between them (Figure 3E). By day 70, the media had a further increase, 72.32% (95% CI 59.84-84.80) for the 25 μg group and 82.66% (95% CI 74.07-91.25) for the 50 μg group, but only significant differences with placebo were found (Figure 3E).

We also measured neutralizing antibody titers against live-SARS-CoV-2 in serum samples of participants with ≥30% of inhibition of RBD-ACE-2 binding. For the short schedule, the proportion of individuals with neutralizing antibodies, at day 42, was 50% for the 25 μg group and 94.73% for the 50 μg group (p=0.029). At day 56, the percentages were 80% for the 25 μg group and 94.7% for the 50 μg group (p=0.55) (Table 2). For the long schedule, by day 56, 100% of individuals had neutralizing antibodies for both 25 μg and 50 μg group, and at day 70, 93.8% and 95.0% for 25 μg and 50 μg group, respectively (p=0.99) (Table 2). For the short schedule GMT values were 18.20 (95% CI 10.72-30.91) for the 25 μg group and 22.80 (95% CI 12.19-42.64) for the 50 μg group (p=0.86) (Figure 3C). At day 56, GMT for the 25 μg group was 10.40 (95% CI 6.83-15.84) and 31.53 (95% CI 19.66-50.59) for the 50 μg group (p<0.01) (Figure 3C). For the long schedule, the GMTs by day 56 were 17.41 (95% CI 2.69-112.50) and 31.09 (95% CI 8.89-108.70) for the 25 μg and 50 μg group, respectively (p=0.37) and at day 70, the 34.63 (95% CI 18.05-66.46) for the 25 μg group and 71.31 [95% CI 38.57-131.83]) for the 50 μg group (p=0.1) (Figure 3C).

In phase 2, blood samples for the evaluation of immune response were taken at time 0 and at days 42 and 56 (14 and 28 days after the third dose). All the results were analyzed globally and stratified by age groups: 19 to 54 years and 55 to 80 years (Table 3). Global seroconversion rates of anti RBD-IgG measured at day 42 were, 79.5% and 89.6% for the 25 μg and 50 μg group, respectively (p=0.00) and at day 56, were very similar with 77.7% and 89.2% for the 25 μg and 50 μg group, respectively (p=0.00). In the age group of 19-54 years, the seroconversion percentages were higher, 87% for 25 μg group and 94.7% for 50 μg group (p=0.04) and, as it was expected, seroconversion rates were lower in the older participants, age 55 to 80 years, 67.7% and 81.3% for the 25 μg and 50 μg group, respectively (p=0.05). The results at day 56 were very similar (Table 3). In the case of placebo group a small number of participants (less than 5%) seroconverted. By day 42 and 56, the global GMT had increased for both 25 and 50 μg groups (39.62 and 93.46) and (36.66 and 80.36) respectively, with higher increases in the 50 μg group and significant difference with the 25 μg group (p<0.0001) (Table 3, Figure 4A). The same results were found for the age group of 19-54 years (with GMTs of 47.84 and 109.86 (p<0.001) and 40.58 and 98.32 (p<0.0001), at 42 and 56 days respectively) (Table 3, Figure 4D). For the oldest group, age 55-80 years, GMTs were of 29.76 and 71.29 (p<0.05) and 27.22 and 57.45 (p<0.05), at 42 and 56 days respectively) (Table 3, Figure 4G). For this age group there were differences between 25 and 50 μg groups (p<0.05) at baseline, although GMTs were very low (Figure 4G).

**Table 3.**
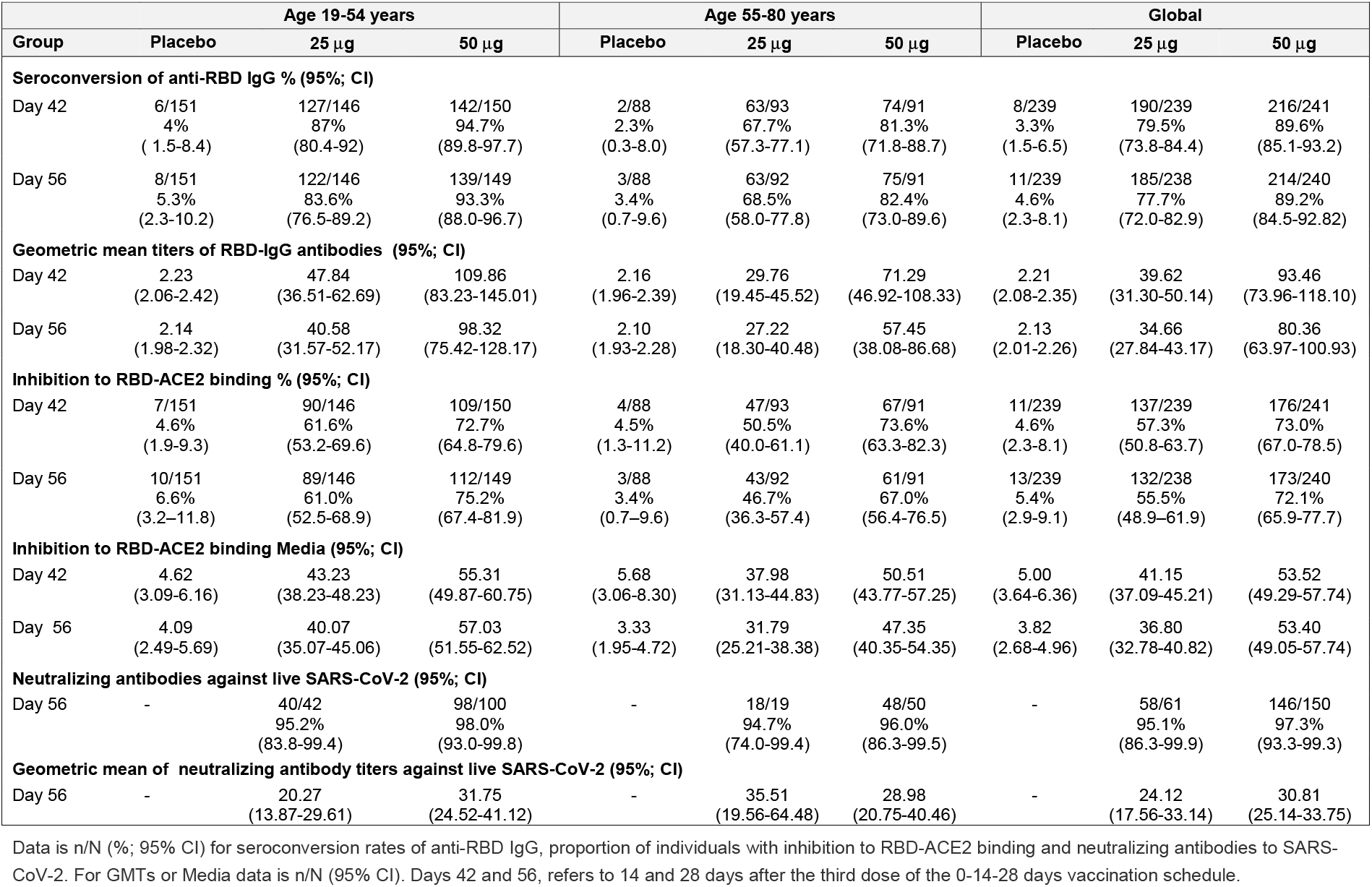
Phase 2 immunological results by study groups, globally and stratified by age (19-54 years and 55-80 years).

**Figure 4.**
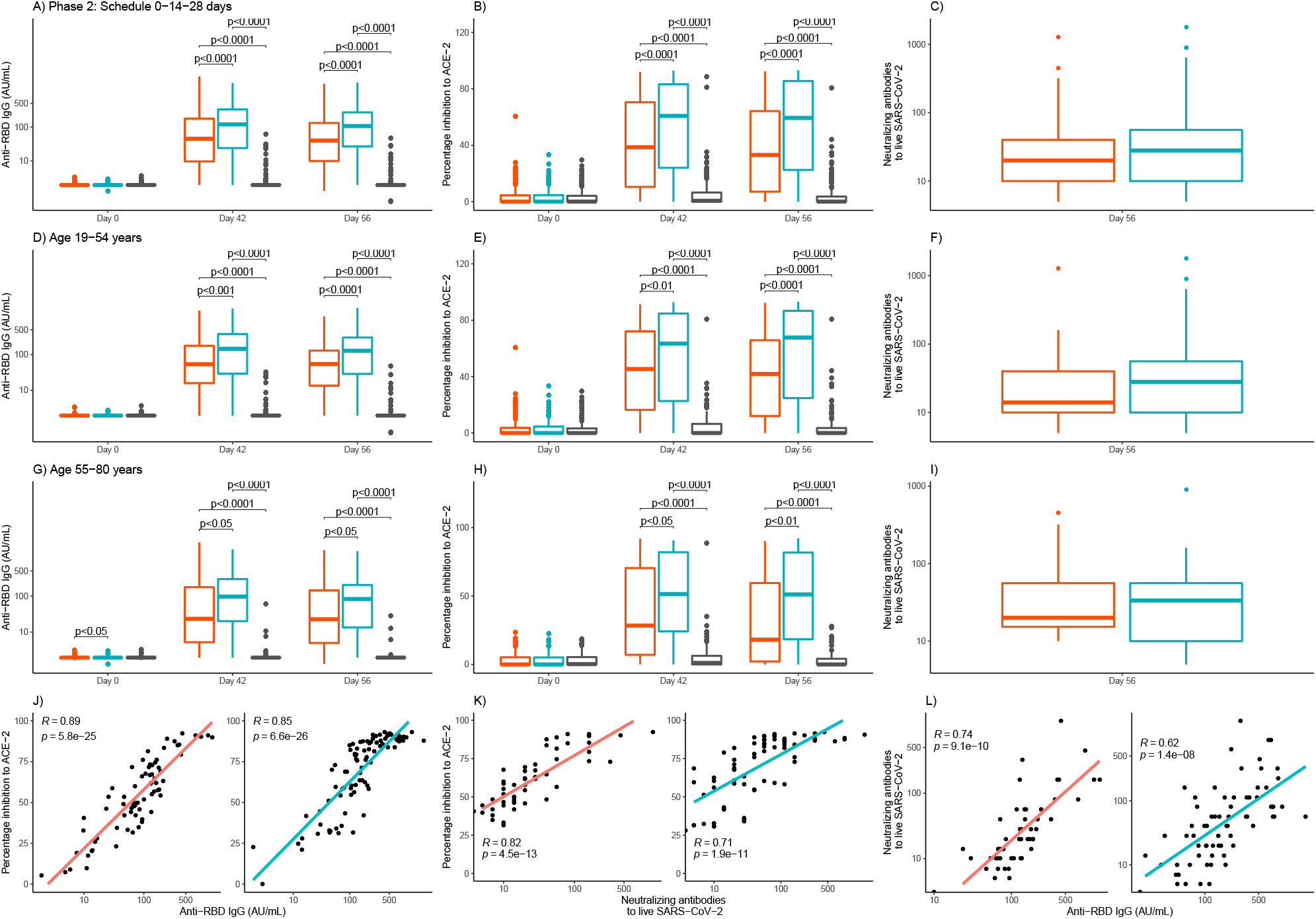
Phase 2 quantitative variables of immunogenicity by study groups and days. From left to right are anti-RBD IgG, percentage of inhibition RBD-ACE2 binding and neutralizing antibody titers, in the global analyses (graphs A, B, C) and stratified by age 19-54 years (graphs D, E, F) and age 55-80 years (graphs G, H and I). The study groups are represented by colors, red for RBD 25 µg, blue for RBD 50 µg and gray for placebo. The boxes and horizontal bars indicate interquartile range (IQR) and the median, respectively. The whisker’s end points are the maximum and minimum values below or above the median ± 1.5 times the IQR. Points represent possible outliers. The braces contain the results of the Mann Whitney U multiple comparison tests with Bonferroni correction. For the viral neutralization variable, Student’s t tests were used to compare geometric means. Only p values for significant differences are shown. The bottom row shows the scatter graphs and the line adjusted by least squares by study group (red for RBD 25 µg, blue for RBD 50 µg) of the values corresponding to day 56. In each case, Pearson’s correlation coefficient and the associated p-value are included.

The proportion of individuals with positive inhibition of RBD-ACE-2 binding at day 42 was of 57.3% and 73.0% for 25 and for 25 μg and 50 μg groups, respectively, (p=0.00) and at day 56 55.5% and 72.1% (p=0.00) (Table 3). The media of the percentage of inhibition of RBD-ACE-2 binding showed an increase at 42 and 56 days, with respect to baseline levels for both 25 μg and 50 μg groups, when analyzed globally or stratified by age groups. At 42 and 56 days, the percentages in the group of 50 μg were significantly higher than the obtained for the 25 μg group (Table 3, Figures 4B, 4E and 4H).

The proportion of individuals with neutralizing antibodies against live-SARS-CoV-2 was measured only at day 56 in a group of participants with ≥ 30% of inhibition of RBD-ACE-2 binding. The results were similar for both 25 and 50 group, with no statistical differences found neither globally nor by the age groups of 19-54 years and 55-80 years (p=0.69, p=0.72 and p=1.00, respectively). GMT of neutralizing antibodies were very similar for 25 μg and 50 μg groups globally (24.12 and 30.81), in the age group of 19-55 years (20.27 and 31.75) and for the age group of 55 to 80 years (35.51 and 28.98) without significant differences between them (p>0.05) (Table 3, Figures 4C, 4F and 4I).

Correlation coefficients were calculated with global data of phase 2 trial, for both 25 μg and 50 μg groups. The correlation coefficient between anti RBD-IgG and the percentage of inhibition to ACE-2 was 0.89 (95% CI 0.82-0.93) for the 25 μg group and 0.85 (95% CI 0.78-0.90) for the 50 μg group (Figure 4J). The correlation coefficient between neutralizing antibodies to live SARS-CoV-2 and the percentage of inhibition RBD-ACE-2 binding was 0.82 (95% CI 0.70-0.89) for the 25 μg group and 0.71 (95% CI 0.56-0.81) for the 50 μg group (Figure 4K). The correlation coefficient between anti RBD-IgG and neutralizing antibodies to live SARS-CoV-2 was 0.74 (95% CI 0.58-0.84) for the 25 μg group and 0.62 (95% CI 0.45-0.75) for the 50 μg group (Figure 4L). Strong and significant correlations were observed in all the cases, for both 25 μg and 50 μg groups (Figures 4J, 4K and 4L).

## Discussion and conclusions

Several vaccine candidates has been developed and found safe and effective against Covid-19. The S protein RBD of SARS-Cov-2 has been the target antigen using different technological platforms such as mRNA, adenovirus vector, inactivated virus and subunit vaccines. [10, 11]

This work reports for the first time the safety and immunogenicity of the Abdala vaccine based in the subunit RBD protein of SARS-CoV-2 in a randomised, double-blind, placebo-controlled trial. The trial performed well, without deviations and a minimum of dropouts. This was facilitated by the fact that vaccination was short lasting and inclusion could be completed easily. Randomisation and blinding assured that bias was minimal. Therefore, the internal validity of the study was adequate.

In phase 1 trial two different immunization schedules were evaluated (0-14-28 and 0-28-56 days). In our approach to develop COVID-19 vaccine it was foreseen that the immunization schedule would be of three doses, based on CIGB’s previous experience with its recombinant hepatitis B vaccine, produced for more than 30 years, whose vaccine antigen is an also a recombinant protein obtained from the same technological platform that uses the SARS-CoV-2 RBD antigen of the Abdala vaccine under study in this trial. This strategy is not the most frequent employed by other vaccine’s developers considering worldwide 15% of the candidates use only one dose and 62% two doses. [11]

The safety and immunogenicity analyses indicated that three doses of Abdala at different strengths and with different immunization schedules in adults 19 to 80 years of age were safe and induced high immune responses, including neutralizing antibodies closely correlated with the anti-RBD IgG response.

The incidence of adverse reactions in the 25 μg and 50 μg group were similar, indicating no dose-related safety. The adverse reactions reported were minimal, mostly mild and from the injection site, of short duration, resolved spontaneously. The most common symptom being injection-site pain, which is in accordance with previous findings for another COVID-19 vaccines. [12-14]

As anticipated, immune responses induced by the 0-28-56 days vaccination schedule were larger than those induced by the 0-14-28 days vaccination schedule, regardless of the dose. However, quick antibody responses could be induced within a relatively short period of time by using a 0-14-28 days schedule. Although it can be argued that the longer vaccination schedule will induce not only a more robust antibody response but also potentially longer persistence of the response could be expected, the actual immune persistence of the two schedules needs to be verified in future studies. However, it was of interest to assess the results on day 56, after applying two doses of the long schedule versus the three doses of the short schedule, in line with the interest to use fewer doses, like other SARS-CoV-2 vaccines based mainly on different technological platforms. In that analysis, for both strengths of the vaccine, significantly higher GMT levels of anti-SARS-CoV-2 IgG antibodies were obtained after three doses of the schedule 0-14-28 days compared to only two doses of the schedule (0-28 days), which reinforced the original idea of the need for three doses of the candidate, under the experimental conditions assessed. Other studies also indicate that a relatively longer interval (21 or 28 days) between injections would be preferred, but the efficacy of the three and two dose schedules is unclear, and the optimal or minimum antibody titers that could protect people form COVID-19 are yet to be established. [15] In phase 1-2 of Coronavac vaccine, quick antibodies response at 0-14 days were obtained but a more robust antibody response was obtained with the 0-28 schedule, although the persistence of the two schedules needs to be verified in future studies. [12]

Considering the capacity to generate a satisfactory immune response with both immunization schedules, including neutralization of the SARS-CoV-2 virus in cell cultures, which is one of the most important variables in relation to the functional capacity of antibodies induced by vaccination and taken into account as a determining practical premise, the exceptional pandemic conditions where it was necessary to immunize people in the shortest possible time, it was decided to continue to phase 2 trial with the three-dose short immunization schedule of 0-14-28 days. This schedule offered a favorable risk-benefit ratio, in terms of safety and immunogenicity. Although most of the COVID-19 vaccines already under emergency authorization are applied mainly in schedules of only two doses, and the three-dose schedule can be considered more complicated and time consuming for massive immunization programs, the short immunization schedule proposed in this trial (0-14-28 days) would allow in only one month to complete the vaccination regimen with good immunogenic profile and potentially a third dose instead of only two doses in this short period of time would be better. This proposal might be also suitable for emergency use, to achieve high vaccination coverage’s in the exposed population of vital importance during the COVID-19 pandemic.

Phase 2 results were consistent with those obtained previously in phase 1 trial, where seroconversion percentages and GMTs of anti-RBD IgG, inhibition percentage of RBD-ACE2 binding neutralizing antibodies confirm the favorable immunogenic profile of the Abdala vaccine with better results and risk-benefit ratio for the 50 μg group. As expected, a better immunological performance of the vaccine was obtained in the age group age 19-54 years that the oldest individuals age 55-80 years. However, for each age stratum and overall for all study participants, the probability of success of each experimental group (RBD 25 and 50 μg) compared to the control (placebo) was estimated, and the hypothesis was fulfilled, since in all cases the probabilities were 1 or values very close to 1. This is demonstrated at both evaluation times, both overall and when stratified by age groups. GMTs of anti-RBD IgG and percentages of inhibition RBD-ACE2 binding also found differences in favor to the 50 μg group but not in the evaluation by day 56 of neutralizing antibodies. It is well-known that there is a large variation in the immune response to vaccination among different individuals, both in quantity and quality, and there is strong evidence that intrinsic factors, such as genetics, sex, age at vaccination, comorbidities, as well as vaccine-related factors (such as choice of vaccine products, adjuvants, and vaccination schedule) strongly influence vaccine responses. [16] Age is considered an important factor influencing the immune response to vaccines, mainly in individuals in the extreme ages of life. This response is thought to be decreased in the earliest stages of life and also in the elderly, who also have a faster decrease in antibodies. This effect has been found for several vaccines where older people have lower antibody levels. [17-24] Taking this knowledge into account; it is not surprising to find lower seroconversion values in individuals aged 55-80 years, even though more than 80% of the subjects seroconverted. Other of COVID-19 vaccines have assess safety and immunogenicity in older individuals showing lower responses when compare with the youngest ages. [25-27]

It should be noted that since most vaccines induce very high antibody responses, small differences in antibody concentrations between groups of individuals may not be clinically significant in terms of protection or efficacy, and may be relevant only in individuals with poor responses, or may affect only the duration of protection, but this has to be proven in phase 3 efficacy trial and the evaluation of the immune response during longer follow-ups. In addition, the quality of the antibody response is an important factor since only a subset of the total detectable antibodies may have functional activity capable of neutralizing pathogens. In this sense, they have more value as potential correlates of efficacy than seroconversion values or geometric mean of titers, which assess the quantity of antibodies but not their functional activity. [16] Moreover, the level of in vitro antibody response does not necessarily correlate with health outcomes, i.e., seroconversion does not mean complete protection against a disease, and non-seroconversion is not necessarily associated with susceptibility, not to mention that antibody levels decline over time, but seronegative individuals may still be protected through other immune mechanisms, as shown, for example, after hepatitis B vaccination. [17]

Neutralizing antibody titer is the most common correlate of protection against viral vaccines and it is highly correlated with protective effect and durability of protection. Results from previous studies on monoclonal antibodies and convalescent sera, as well as tests in animal models, have all confirmed the role of neutralizing antibodies in conferring protection against COVID-19. [28, 29] Neutralizing antibody responses to SARS-CoV-2 in hospitalized COVID-19 and convalescent patients are above 160 in more than 93% of convalescent sera. [30, 31] However, in the different clinical trials performed the GMT of the neutralizing antibodies varies for the different vaccines. In addition, comparisons between the different vaccines developed may not be reliable and indeed not be comparable, due to the lack of standards that could serve as reference points in the studies, and the different methods used to assess this response. [32]

Finally, when Pearson’s linear correlation analyses were performed between the different immunogenicity variables, positive and highly significant correlations were found, as described in the corresponding results section, demonstrating the relationship that existed between these variables, indicating that the immune response for the vaccine is not only potent in quantity but also in the quality of the antibodies elicited.

The lack of standards and use of different assays complicate the comparison of performance of the various COVID-19 vaccines and the comparisons of convalescent serum panels could be rather arbitrary, taking into account the variations in the composition of the panels. Given varied assays across clinical studies, it was also inappropriate to directly compare antibody titers in our study with other trials. Very recently blood samples from individuals vaccinated with Abdala vaccine were evaluated using Elecsys^®^ Anti-SARS-CoV-2 S (Roche Diagnostics) and showed a high correlation between the Cuban tests UMELISA SARS-CoV-2 ANTI RBD (used in this phase 1-2 trial) and Roche’s Elecsys^®^ test, demonstrating that our vaccine reached antibody titers comparable to those of Pfizer/BionTech vaccine using the same test. [33]

This study has several limitations. First, immunogenicity was tested at day 14 and 30 after complete vaccination schedule, so the duration of the immune response cannot be assessed. Follow-up visits to evaluate long-term safety as well as the duration of the immune response at least 6 months after vaccination are underway. Second, we did not assess the T cell responses in the phase 2 trial. Third, the relevance of antibody response elicited by this vaccine to protection against COVID-19 disease has to be evaluated in phase 3 efficacy trials. Four, data of neutralizing antibody titers against emerging variants of SARS-CoV-2 require further studies currently ongoing.

The ethnic diversity of the Cuban population, the wide age range studied and its comorbidities, contributes to the generalisability (external validity, applicability) of the trial findings, although they are still limited the sample size.

In conclusion, the results of the phase 1-2 trial indicated that Abdala vaccine against SARS-CoV-2 was safe, well tolerated and induced humoral immune responses against SARS-CoV-2 among adults from 19 to 80 years of age. Our findings indicate that the a SARS-CoV-2 recombinant spike protein vaccine studied (Abdala) is a promising candidate that warrants testing in phase 3 studies, in a larger number of individuals older than 19 years of age and a three-dose schedule of 50 μg on days 0-14-28, evaluating vaccine efficacy in the prevention of symptomatic COVID-19 and progression to serious and critical forms of the disease.

## Data Availability

All data produced are available at data base, clinic history, Cuban drug regulatory agency, ecc.

## Data Availability

All data produced are available at data base, clinic history, Cuban drug regulatory agency, ecc.

## Acknowledgements

The authors wish to acknowledge the “Saturnino Lora” Hospital Direction and the Public Health Ministry of Cuba for its support.

## Conflict of Interest Disclosure

Authors FHB, YMB, JQG, KUP, JLRR, MAV, MLF, GEGN, GLP, MAA, and VLMG, are employees of the Center for Genetic Engineering and Biotechnology, Havana Network, where Abdala vaccine active ingredient is produced and the formulation was developed. The remaining authors have no conflict of interests. No honoraria, consulting fees or payments for seminar presentations, speeches or appearances have been received by any of the authors. The study was financed by CIGB, Havana (products, reagents) and the Ministry of Public Health of Cuba (hospital facilities and general medical care of the participants).

## Appendix

**Other Investigators: “Saturnino Lora” Hospital, Santiago de Cuba:** Yaquelín Naranjo-Vargas, Ahimara Rosado-Rosado, Rafael Suárez-Domínguez, Liudmila R. Castro-Andión, Mercedes Gay-Muguercia, Miguel A. Vinent-Terazón, Liader Aguilera-Ramos, Lisbeth Mantejo-Hormigó, Daniela Ramis-Rosales, Dariana Ávila-Fuentes, Mari C. Acosta-Portuondo, Héctor Pérez-Hernández, Yurina Álvarez-Calvo, Virgen Ruano-González, Luisa Recacén-Canet, Odalis Poll-Fiss, Sandra Jaime-Vázquez, Yaima Ferrer-Bonne, Eneida Sánchez-Carbonell, Yarelis Sotomayor-Murgada, Ledys Garbey-Delás, Juliet Sigüenza-Castilla, Magda Domínguez-Cardosa, María Clares-Pochet, Ofelia Veliz-Aranda, Yordenis Días-Hodelín, Irma Ramírez-Bicet, Sara Hernández-Falls, Obadis Nordet-Figueras, Rosa Ferrer-Socarrás, Elizabeth Ramos-Caraballo, Gilberto Moya-Justiz, Odalexis Arias-Ramos, Lixania Tamé-Aguilar, Damaris Veranes-Fernández, Gloria Sánchez-Cintra, LIilian Velázquez-Zayas, Esther Mayor-Guerra; **Center for Genetic Engineering and Biotechnology:** Elizeth García-Iglesias, Karem Catasús-Álvarez, Grettel Melo-Suárez, Ricardo Martínez-Rosales, Sheila Chávez-Valdés, Lismary Ávila-Díaz, Ana Campal-Espinosa, Edelgis Coizeau-Rodríguez, Yahima Chacón-Quintero, Giselle Freyre-Corrales, Amalia Vázquez-Arteaga, Hany González-Formental, Franklin Aguilar-Fuentes, Mónica Bequet-Romero; **Institute of Cybernetics, Mathematics and Physics, Havana:** Jesús E. Sánchez-García, Ernesto Rodríguez-Martínez, Rolando Selgas Lizano; **Biotechnology and Pharmaceutical Industries Group, BioCubaFarma, Havana:** Eulogio Pimentel-Vázquez, Eduardo Martínez-Díaz.

## Notes

### Clinical Trial

RPCEC00000346. Cuban Public Clinical Trial Registry (WHO accepted Primary Registry).

### Author Declarations

Ethics committee/IRB of Provincial Hospital "Saturnino Lora" in Santiago de Cuba, gave ethical approval for this work.

